# Deciphering Physiological and Pathological Influences on Amygdala-Hippocampus Connectivity

**DOI:** 10.64898/2026.09.06.26362367

**Authors:** Odile Feys, Mariam Josyula, Nishant Sinha, Marc Jaskir, Samuel B. Tomlinson, Caren Armstrong, Sandhitsu Das, Joel M. Stein, Eric D. Marsh, Kathryn A. Davis, Erin C. Conrad

**Author notes:** Equal contribution (Kathryn A. Davis, Erin C. Conrad). Corresponding authors: Erin C. Conrad and Odile Feys, Department of Neurology, Perelman School of Medicine, University of Pennsylvania, 210 South 33^rd^ Street, 304 Hayden Hall, Philadelphia, /.

## Abstract

**Objective:** Temporal lobe epilepsy (TLE) is associated with disrupted functional integrity in the amygdala-hippocampus complex. Cortico-cortical evoked potentials (CCEPs) can characterize this disruption and have been proposed as biomarkers of the epileptogenic zone (EZ), but their study is typically limited by the spatial sampling bias inherent to whole-brain intracranial EEG. We investigated how epileptogenicity shapes effective connectivity in the amygdala-hippocampus complex, whether structural connectivity underlies it, and ultimately derived a multimodal EZ biomarker.

**Methods:** We retrospectively included 71 patients (50 adults, 21 children) who underwent single-pulse electrical stimulation protocols with intracranial contacts in the amygdala or hippocampus; 15 also underwent diffusion MRI. CCEPs were visually detected, and the latency and amplitude of the first response peak (D1) were extracted. Structural connectivity metrics (tract length, quantitative and fractional anisotropy, mean diffusivity) were derived between the same contacts. A Bayesian linear mixed model (BLMM) related D1 latency to clinical, neurophysiological, and structural predictors, handling missing DTI values jointly within the model. A corrected latency score was then built to discriminate epileptogenic from non-epileptogenic contacts.

**Results:** Among 6257 possible stimulation-recording pairs, 1027 CCEPs were detected with a significantly higher rate in the hippocampus compared to the amygdala. The BLMM identified robust associations between D1 latency and epileptogenicity, epilepsy type, ipsilateral stimulation, stimulation site (hippocampus/amygdala), and quantitative and fractional anisotropy. The resulting EZ score, obtained by extracting the EZ term’s contribution from the BLMM equation, demonstrated an ability to discriminate epileptogenic contacts, showing a balanced accuracy of 78% (sensitivity 86%, specificity 71%), and the resulting EZ probability, based on an elastic net logistic regression, showed a balanced accuracy of 84% (sensitivity 86%, specificity 82%).

**Discussion:** These findings suggest that effective connectivity results from the interplay of opposing physiological (here amygdala vs. hippocampus) and pathological (epilepsy-related) influences rather than a simple facilitation within the EZ, and that white matter microstructure independently contributes to this timing. Connectivity is slower within the EZ itself, with an even greater delay observed in its vicinity compared to other brain areas. The resulting EZ score offers a practical, closed-form tool to strengthen EZ localization, and paves the way toward a structurally informed, CCEP-based framework extendable to other brain regions.

## Introduction

Temporal lobe epilepsy (TLE; i.e., the most common type of human focal epilepsy, frequently referred for epilepsy surgery (1)) is characterized by a disruption of the functional organization of the amygdala-hippocampus complex (2), a structure considered central to the epileptogenic network in this condition (3). Characterizing how the connectivity of this complex differs between epileptogenic and non-epileptogenic contacts would improve our understanding of TLE and may allow us to better target epilepsy surgery.

Cortico-cortical evoked potentials (CCEPs) represent a measure of the effective connectivity (i.e., directed functional connectivity (4)) between a brain area where an intracranial stimulation (single-pulse electrical stimulation (SPES)) is performed and the brain area where its response can be recorded (5), and have been studied in recent years as potential biomarkers of the EZ, with several of their features related to their localization inside or outside the EZ (6–12). Their interpretation, however, is typically complicated by the spatial sampling bias inherent to intracranial EEG, most commonly stereo-EEG (SEEG), which investigates only around 0.5% of the total brain volume (13), and by the fact that implantation schemes differ between patients depending on patient-specific characteristics, e.g., brain anatomy or seizure semiology (14). To overcome spatial sampling bias, studies have also combined intra- and extracranial recordings to study CCEPs at the whole-brain level (15,16), demonstrating that, beyond EZ and distance between areas, intrinsic physiological characteristics of anatomical regions also impact CCEP features (16,17). Nonetheless, these simultaneous recordings remain rare. Focusing on the amygdala-hippocampus complex, a brain area that is both highly and consistently implanted during SEEG recordings (18), mitigates both biases: spatial sampling bias is greatly reduced, and region-specific connectivity is controlled, since the same anatomical region is investigated across all patients.

Still, CCEPs characterize only the functional response to stimulation and do not reveal the anatomical pathways that support this propagation. Determining whether variations in CCEPs reflect the structural wiring itself, or an effect of pathology independent of this wiring, requires an additional measure of structural connectivity, e.g., derived from diffusion tensor imaging (DTI) (19). This multimodal analysis combining functional and structural connectivity may be applied to the study of CCEPs (20) (Figure 1), and was performed here in a subgroup of patients with available diffusion imaging data.

**Figure 1.**
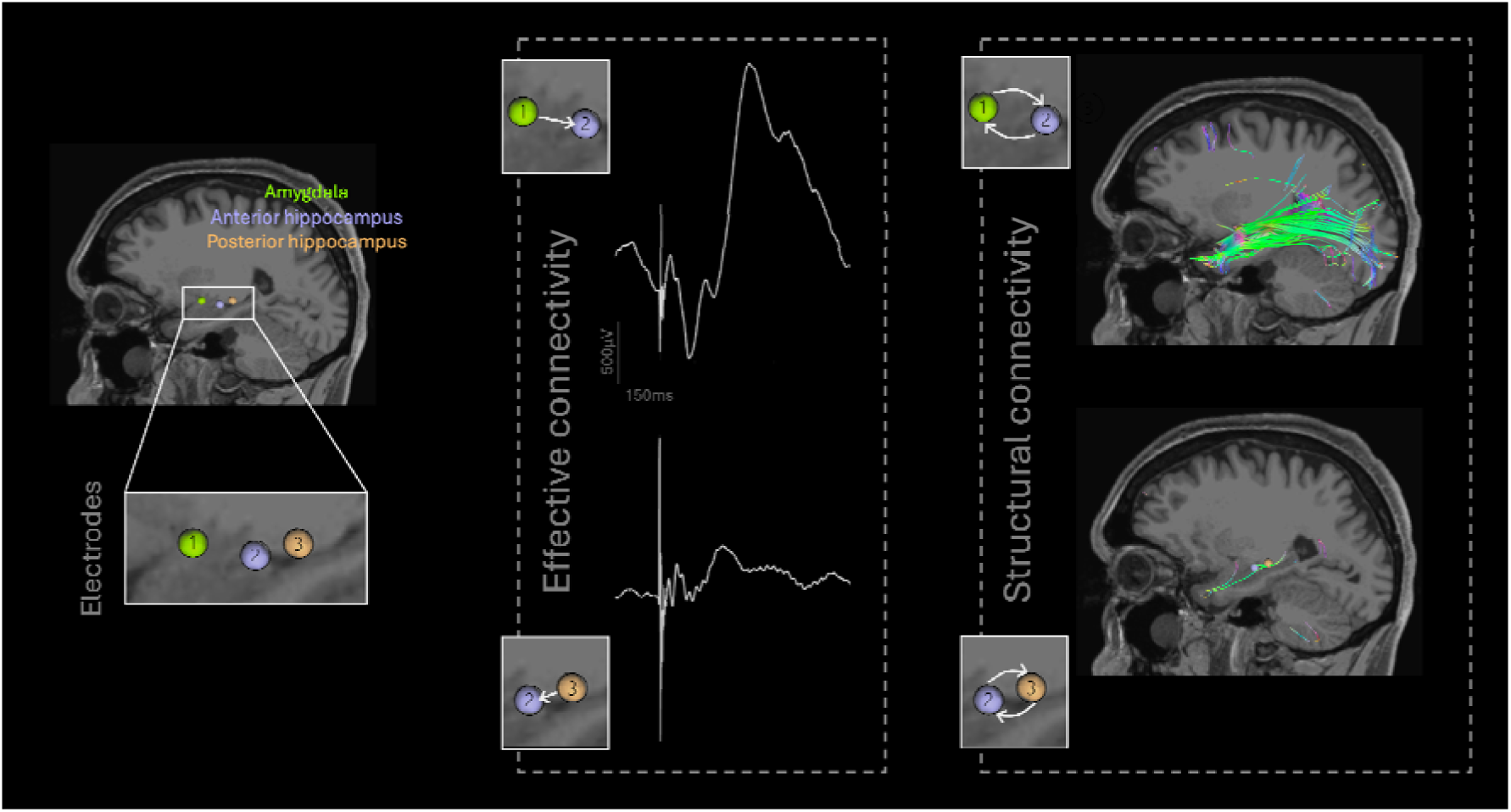
Effective and structural connectivity in the amygdala-hippocampus complex. **Left.** Intracranial contacts in the amygdala (1, green), anterior hippocampus (2, lavender), posterior hippocampus (3, orange) of an example patient. **Middle.** Two CCEPs recorded in the anterior hippocampus of this patient, one following amygdala stimulation (top) and one following posterior hippocampus stimulation (bottom; identical scale). CCEPs measure effective connectivity, also named directional functional connectivity, comprising the direction of the connectivity (i.e., region A influencing region B or region B influencing region A are discriminated). **Right.** White matter tracts passing through both amygdala and anterior hippocampus (top) and anterior and posterior hippocampus (bottom), no more than 3mm apart from the studied intracranial contacts. Diffusion MRI measures structural connectivity which does not indicate the direction of the connectivity (i.e., which of regions A or B influence the other region is not known).

This study aims to investigate (i) the specific effect of epileptogenicity on effective connectivity by focusing on a very precise brain area (i.e., amygdala-hippocampus complex), (ii) the underlying effect of structural connectivity on effective connectivity, (iii) the classification of brain regions as epileptogenic vs. non-epileptogenic based on brain connectivity.

## Methods

### Inclusion criteria

We retrospectively included all patients who underwent SPES protocols at Hospital of the University of Pennsylvania (HUP) and Children’s Hospital of Philadelphia (CHOP). At CHOP, SPES were performed under research protocol IRB15012226. At HUP, SPES were performed both as part of a research protocol under IRB821778 and as part of routine clinical care. In both cases, patients consented to retrospective analysis and data sharing. SEEG was performed as part of routine presurgical assessment following a noninvasive evaluation (including at least medical history, neurological examination, neuropsychological testing, scalp video-EEG, MRI). The implantation scheme was determined during the surgical meeting discussion based on the noninvasive evaluation.

### SEEG recording, localization, stimulation

Surgical implantation of multiple depth electrodes (Ad Tech Medical Instruments; 6 to 12 intracranial contacts, contact length: 2.41mm, contact diameter: 1.1mm, intercontact spacing: 5mm) was performed using a robot-assisted procedure. Contact localization at HUP was achieved through in-house software applied to linearly registered preimplantation and postimplantation T1-weighted brain MRI and postimplantation brain CT, as previously described in (21,22); at CHOP, it was performed using Gardel software (23). Intracranial signals were acquired using a Natus Quantum amplifier (sampling rate: 512 to 2048Hz, anti-aliasing filters: 219-439-879Hz). The SPES protocol consisted of bipolar stimulation delivered via a Nicolet Cortical Stimulator (Natus) with the following parameters: amplitude 3mA, pulse width 300-500μs, frequency 1Hz, train duration 30s at HUP and 1-8 mA, 300-500μs, 1-2 Hz, 30s at CHOP. Intracranial data were then extracted, referenced in bipolar montage, and averaged following stimulation artifact detection. Further preprocessing details are provided in (8).

### Analysis of amygdala-hippocampus CCEPs

Intracranial contacts located in the amygdala or hippocampus were selected, and a notch filter (60Hz and harmonics) was applied. Given the risk of false positives inherent to automated approaches, visual detection of CCEPs was performed (24). Each stimulating-recording electrode pair was visually inspected by O.F., blinded to the epilepsy localization, to exclude those presenting: (i) residual powerline noise, (ii) cerebral noise (e.g., continuous slow waves or intermittent spikes), or (iii) absence of evoked potentials (Figure 2, middle). For electrode pairs with visually confirmed CCEPs, D1 (defined as the first peak, positive or negative, following the stimulation artifact (7), also referred to as N1 in the literature (25)) was visually identified by O.F. (Figure 2, right; after check on D1 detection concordance between two independent observers, O.F. and E.C.C.), and its timing and amplitude were subsequently extracted.

**Figure 2.**
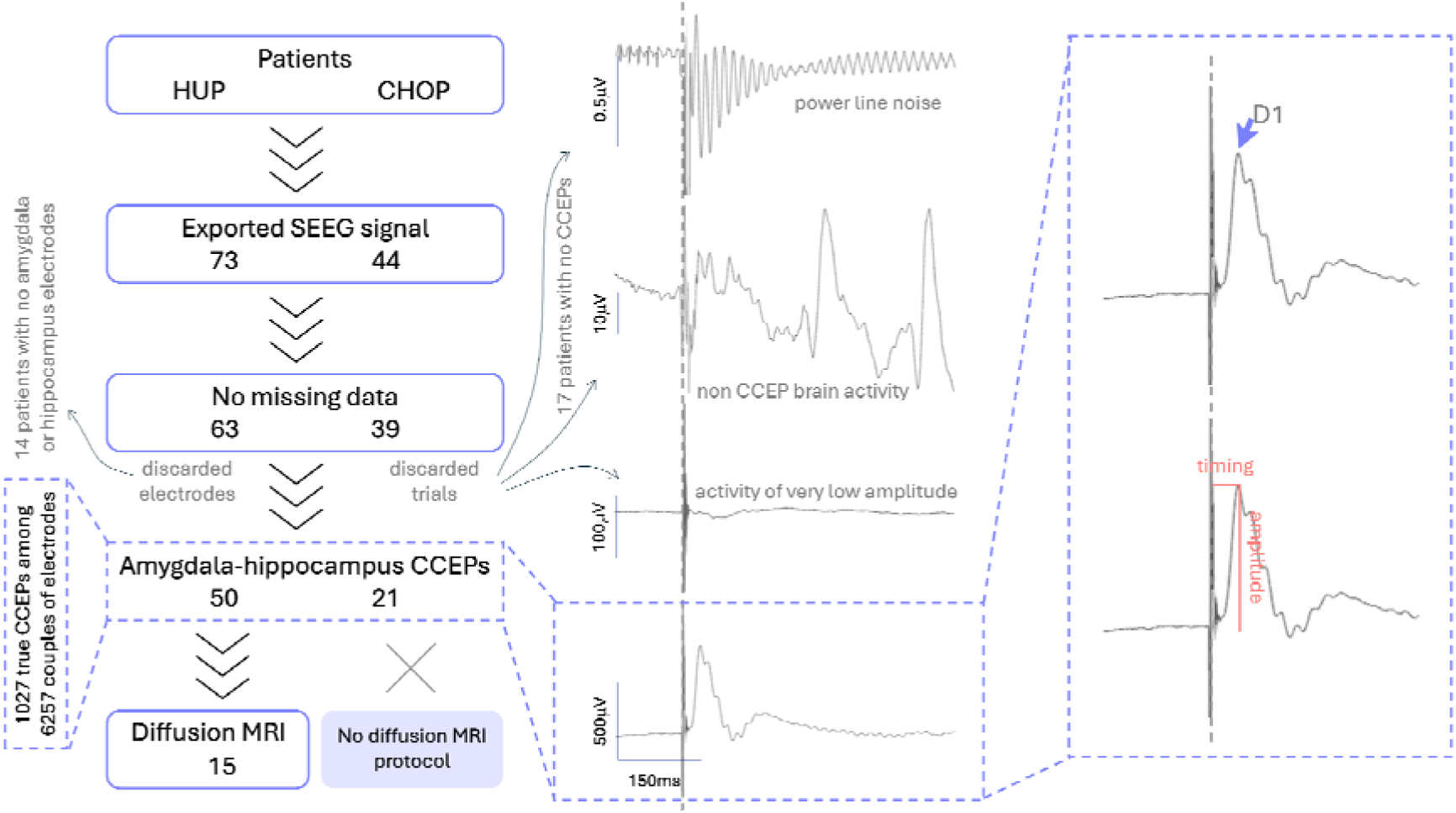
Patients’ selection and CCEPs detection. Across the two included centers, 117 patients underwent SPES protocols (73 adults and 44 children). Fifteen patients had missing data, including non-delineated EZ during SEEG which forces the rejection of these patients for further analyses. Fourteen patients had no electrodes in the amygdala or hippocampus, due to the clinically oriented implantation scheme, and are a fortiori rejected. Seventeen patients had at least one electrode, one of them with only one contact in the amygdala-hippocampus complex impeding simultaneous stimulation and recording in the same brain area and the 16 remaining patients showed power line noise, non CCEP brain activity (e.g., spikes or slow waves) or activity of very low amplitude non compatible with CCEP. Among the 71 patients with detected CCEPs, 1027 CCEPs were detected from 6257 possible trials, latency of the first peak (D1) was manually determined, and amplitude was interpolated. Fifteen out of the 50 adult patients with detected CCEPs also underwent a diffusion MRI research protocol.

### MRI acquisition and processing

Preimplantation and postimplantation T1-weighted brain MRI (3T Siemens Prisma scanner, sagittal 208-slice MPRAGE sequence, TE/TR = 2.24/2400ms, TI = 1060ms, FOV = 256mm, 0.8 isotropic voxel size) and preimplantation diffusion-weighted brain MRI (single-shot echo planar imaging multi-shell sequence, 116 diffusion sampling directions, b-values: 0, 300, 700, and 2000 s/mm², resolution: 2 × 2 × 2 mm³, FOV = 220 mm, TR = 4.3 ms, TE = 75 ms) were used to compute patient-specific structural brain networks between depth electrodes. B0 inhomogeneities in diffusion-weighted imaging (DWI) acquisition were corrected using top-up sequences with reverse phase encoding directions or B0 phase and magnitude images. Generalized q-sampling imaging reconstruction was then applied in DSI Studio (diffusion sampling length ratio: 1.25), followed by deterministic tractography (26), yielding approximately 2 million streamlines per subject. Tractography parameters were as follows: Runge-Kutta method, step size of 1 mm, whole-brain seeding, all fiber orientations as initial propagation direction, minimum streamline length of 15mm, maximum streamline length of 300mm.

### Structural connectivity based on DTI

Diffusion tensor imaging (DTI) metrics were derived from the diffusion tensor computed in each voxel (as described in (27)), structural brain connectivity were computed between depth electrodes previously selected in the amygdala or hippocampus. For each electrode pair, connectivity matrices were derived for the following parameters: tract length, quantitative anisotropy (QA), fractional anisotropy (FA), mean diffusivity (MD), diffusivity along the axonal axis (AD) and diffusivity perpendicular to the main axis (RD). FA quantifies the degree to which water diffusion deviates from isotropy: it ranges from 0, when diffusion is unrestricted and equal in all directions, to 1, when diffusion is restricted to a single dominant direction, as is typically the case along coherently organized axons (28). MD represents the average magnitude of water diffusion across all directions, independent of directionality, and is commonly used as a marker of overall tissue microstructural integrity (28). AD refers specifically to the magnitude of diffusion along the principal (longitudinal) axis of a fiber bundle, while RD refers to the magnitude of diffusion perpendicular to that axis; decreased AD have been associated with axonal damage and increased RD with myelin damage (29). QA is derived from the spin distribution function rather than from the DTI and estimates the density of water spins diffusing along a specific fiber direction after subtracting the isotropic (non-directional) background signal (30).

### Clinical data collection

Clinical data were retrospectively collected from medical files of patients including age at the time of SEEG recording, age at epilepsy onset, sex, type of epilepsy (TLE vs. ETLE) and subtype (mTLE vs. other), left vs. right vs. bilateral EZ (defined as the area of the beginning and initial organization of seizures (31)), concordant lesion with EZ, hippocampal sclerosis or not, resective surgery vs. LITT vs. no curative treatment, postoperative Engel class at one year. The type of epilepsy was determined based on the consensus of a multidisciplinary clinical conference incorporating the results of SEEG and pre-implant data. TLE includes all patients with medial or lateral temporal lobe involvement in the EZ and mTLE includes all patients whom the amygdala or hippocampus were considered in the EZ.

### Statistical analyses

#### D1 detection between independent observers

Agreement between raters on the presence of an early response (D1 < 70 ms (25)) was quantified as the percentage of 1000 random trials with concordant classification. Among trials for which both raters identified an early peak, latencies were compared using a two-tailed Wilcoxon signed-rank test and their association was assessed with Spearman’s rank correlation.

#### Amygdala-hippocampus connectivity

Contingency table was used to compare amygdala vs. hippocampus CCEPs evoked by amygdala vs. hippocampus stimulation using Fisher exact test.

#### Collinearity checks

Point-biserial correlation was used to check collinearity between age and hospital, due to inclusion of pediatric patients from CHOP and adult patients from HUP: if non-collinear, hospital (HUP vs. CHOP) will be considered as categorical variable in addition to age as continuous variable, if collinear (correlation >0.7 (32)), age will be preferred to hospital due to its continuous status which could enrich the model.

Spearman correlation was used to check collinearity between continuous MRI tract metrics (tract length, QA, FA, MD, AD, RD): if two variables showed a correlation coefficient above 0.7 in absolute value, only one of the pair would be retained in the model to avoid redundancy.

#### Models based on neurophysiological variables

None of the patient or observation has a missing value for the neurophysiological variables (i.e., due to the recruitment of this study, each observation is related to a D1 peak whose timing and amplitude can be studied). We first built linear mixed models (LMM) with (i) timing and (ii) amplitude of D1 as outcome variables. For that purpose, we explored all combinations of predictors (see Supplementary Materials) and set subjects as a random variable in all the formulas. We selected the model with the lowest Bayesian information criterion (BIC) as the most parsimonious model, which penalizes model complexity and was used to identify the most supported model among candidate models, and considered models with a ΔBIC<2 as statistically equivalent (33). Among equivalent models, we retained those with a significant global F-test with a two-sided maximum statistic (34) with 2,000 label permutations (35) to correct for multiple comparisons with no independence assumption (significance: p<0.05) and inspected individual predictors via post-hoc marginal analysis of variance (ANOVA) with Satterthwaite approximation for degrees of freedom. Homoscedasticity was checked using residual plots, and models using the logarithm of the outcome variable were built when heteroscedasticity was detected.

#### Confirmation in patients with favorable outcome

In patients with favorable outcome (Engel 1 at one-year post-resection), we performed a secondary analysis re-testing the variables previously identified as significant for D1 amplitude/timing in the models above (significance assessed with global F-test: p<0.05).

#### Stratification of different brain areas

We performed stratified analyses on amplitude/timing of D1 following hippocampus vs. amygdala stimulation using variables demonstrated significant in the models above, to disentangle plausible physiological effect between two brain areas (significance assessed with global F-test: p<0.05).

#### Model based neuroimaging and neurophysiological data

To examine the effect of structural connectivity on CCEPs while accounting for the partial availability of DTI data, we fitted a Bayesian linear mixed model (BLMM) to explain the relevant outcome variable(s) of D1 (amplitude and/or timing depending on the results of models tested on neurophysiological variables). For that purpose, we explored one model including all the predictors (Figure 3; see Supplementary Materials) and set subjects as a random variable.

**Figure 3.**
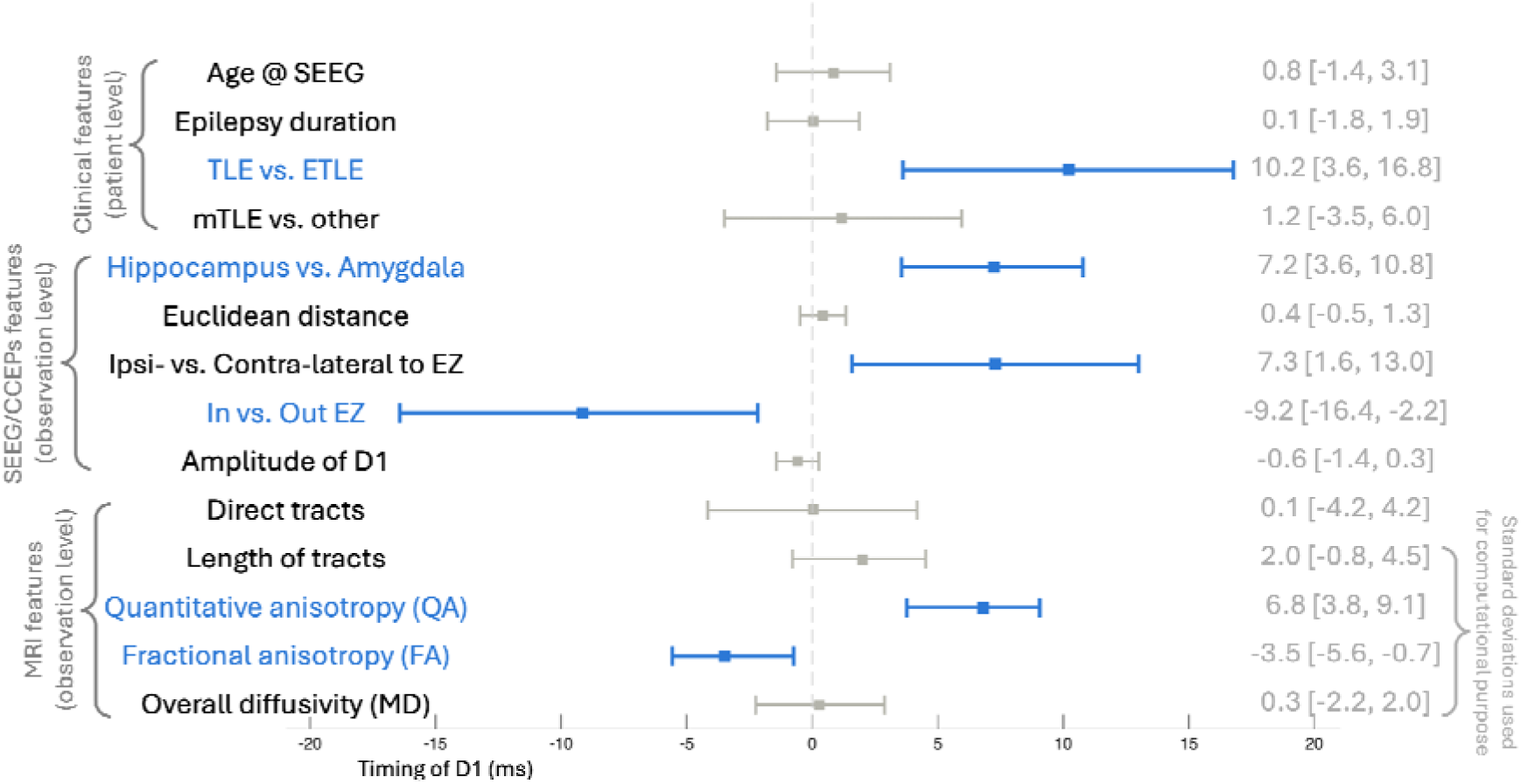
Forest plot of the Bayesian linear mixed model. Predictors are grouped by level of analysis: clinical features recorded at the patient level (top), SEEG/CCEPs features recorded at the observation level (middle), and MRI features recorded at the observation level (bottom). For each predictor, the posterior mean estimate and its 95% credible interval are displayed. Predictors shown in blue indicate robust effects, defined as credible intervals not crossing zero. Predictors shown in grey did not reach this criterion.

Missing DTI values were handled directly within the Bayesian model, which treats missing observations as unknown parameters estimated jointly with all other model parameters. This approach avoids both removal of observations with missing DTI values and multiple imputation, and propagates uncertainty about missing values into the posterior distributions of the regression coefficients. Sub-models for each partially observed DTI variable included significant variables resulting from models on neurophysiological variables as predictors (considered the most plausible determinants). All continuous variables were standardized prior to model fitting to improve sampler efficiency and comparability of effect sizes. Weakly informative priors were used for all parameters (36), see Supplementary Materials.

The model was fitted using the No-U-Turn Sampler (37) with 4 chains of 8000 iterations each (4000 warmup), run in parallel. Convergence was assessed using the potential scale reduction factor (RL < 1.01) and bulk and tail effective sample sizes. Effects were considered robust when the 95% credible interval (CI) of the posterior distribution did not include zero (i.e., Bayesian analogue of statistical significance). The Bayesian R² for the outcome model was computed as a measure of explained variance.

#### Testing complementary information between CCEPs and DTI metrics

To test whether significant DTI metrics and CCEPs features provide complementary information for classifying the EZ, we restricted the analysis to patients/observations with both modalities available. We fitted mixed-effects logistic regression models with a patient-specific random intercept, comparing a reduced model containing the “nuisance” modality with a full model additionally including the modality of interest (i.e., DTI metrics alone versus DTI metrics + CCEPs features, and CCEPs features alone versus CCEPs features + DTI metrics), significance was assessed using a permutation-based likelihood-ratio test adapted from the Freedman-Lane approach, see Supplementary Materials.

#### Building a biomarker of EZ

To evaluate the ability of a BLMM-derived score to discriminate epileptogenic from non-epileptogenic contacts in the amygdala-hippocampus complex, a corrected latency score was computed for each recording contact with available DTI data. Specifically, the D1 latency was adjusted by removing the contributions of all predictors except EZ membership. After isolating EZ in the equation, its sign was inverted so that higher scores correspond to epileptogenic contacts. Receiver operating characteristic (ROC) analysis was performed using this score against the binary EZ label, and the area under the curve (AUC) was computed. Statistical significance was assessed using a permutation test (n = 10,000 permutations) in which EZ labels were shuffled independently within each patient, preserving inter-patient variance. The p-value was defined as the proportion of permuted AUCs showing an effect size (|AUC − 0.5|) equal to or greater than the observed one. The optimal classification threshold was determined by the Youden index (maximizing sensitivity + specificity − 1). Confidence intervals (95%) for the AUC, sensitivity, specificity, and optimal threshold were estimated by patient-level bootstrap resampling (n = 10,000 iterations), ensuring that all contacts from a given patient were resampled together to account for within-patient correlation. Model stability was further assessed using leave-one-patient-out cross-validation (see Supplementary Materials).

To evaluate whether EZ status could be discriminated using neurophysiological and neuroimaging data alone, independent of clinical assumptions embedded in the BLMM, a second classification approach was developed. Because a continuous corrected-latency score analogous to the one described above could not be meaningfully constructed or oriented without incorporating these clinical covariates, EZ probability was instead estimated directly using a supervised machine-learning approach. An elastic net logistic regression (mixing parameter α = 0.5) was fitted with significant neurophysiological/neuroimaging data and a random intercept for patient, using the restricted subset of contacts with available DTI data. The regularization path was estimated via 10-fold cross-validation over a grid of 100 candidate λ values, and the λ minimizing cross-validated deviance (λ_min) was retained (see results for λ_1SE in Supplementary Materials). Model performance was assessed using leave-one-patient-out cross-validation restricted to patients with at least one DTI-available contact: at each iteration, the elastic net model was refit on all remaining patients (with predictor standardization recomputed on the training set), and the resulting coefficients, together with a Youden-optimal threshold determined on the training folds, were applied to the held-out patient’s contacts. Classification metrics (balanced accuracy, sensitivity, specificity, accuracy) were pooled across held-out patients, and 95% confidence intervals were obtained via patient-level bootstrap resampling (n = 10,000 iterations), resampling all contacts from a given patient together.

To exclude clinical information being the most important factors in these models, we tested a null model (see Supplementary Materials).

## Results

### Demographic data

Fifty adults from HUP (mean age: 38.3y, range: 21-65y, 22M/28F) and 21 children from CHOP (mean age: 11.9y, range: 4-18y, 12M/9F) were included in this study (Table 1) from the 117 patients who underwent SPES protocol (Figure 2, left). Twelve patients had bilateral implantation of the amygdala-hippocampus complex, 67 patients had implanted contacts in the hippocampus and 57 patients in the amygdala. Fifteen adult patients from HUP underwent the diffusion MRI research protocol including multi-shell diffusion MRI.

**Table 1.** Demographic and clinical data.

| **Variables** | **Total** | **HUP** | **CHOP** |
| --- | --- | --- | --- |
| Age at SEEG (mean ± standard deviation, years) | 30.5 ± 16 | 38.3 ± 12 | 11.9 ± 4.9 |
| Age at epilepsy onset (mean ± standard deviation, years) | 18.5 ± 15.5 | 23 ± 14.8 | 7.7 ± 4.9 |
| Epilepsy duration (mean ± standard deviation, years) | 12 ± 12.2 | 15.3 ± 13.1 | 4.2 ± 3.5 |
| Epilepsy type (n patients) |  |  |  |
| ETLE | n=14 | n=8 | n=6 |
| TLE | n=57 | n=42 | n=15 |
| *Including mTLE* | *n=39* | *n=31* | *n=8* |
| EZ lateralization (n patients) |  |  |  |
| Left | n=37 | n=26 | n=11 |
| Right | n=26 | n=17 | n=9 |
| Bilateral | n=8 | n=8 | n=1 |
| Structural lesion (n patients) | n=29 | n=16 | n=13 |
| *Including hippocampal sclerosis* | *n=9* | *n=8* | *n=1* |
| Surgery (n patients) | n=39 | n=28 | n=11 |
| *Including patients with one-year outcome* | *n=31* | *n=20* | *n=11* |
| Postoperative outcome at one year |  |  |  |
| Engel 1 | n=23 | n=13 | n=10 |
| Engel 2 | n=5 | n=5 | n=0 |
| Engel 3 | n=1 | n=3 | n=0 |
| Engel 4 | n=2 | n=4 | n=1 |

### Descriptive report of cortico-cortical evoked potentials

Among the 6257 combinations of intracranial contacts compatible with stimulation and recording within the amygdala-hippocampus complex, 1027 CCEPs were detected (Figure 2, left): 678 following hippocampus stimulation and 349 following amygdala stimulation.

O.F. and E.C.C. agreed on the presence of D1 in 84% of a random set of 1000 trials. Among trials with concordant early detection, peak latency did not differ significantly between raters (41.9 ± 9.8 ms vs. 43.9 ± 10.7 ms; Wilcoxon signed-rank test, p = 0.12) and was strongly correlated (ρ = 0.77, p < 0.001).

### Amygdala-hippocampus connectivity

The hippocampus-amygdala connectivity (0.1%, defined by the existence of a CCEPs between two contacts) and amygdala-amygdala connectivity (0.4%) were significantly lower than amygdala-hippocampus connectivity (33.6%) and hippocampus-hippocampus connectivity (65.9%; Fisher test, p=0.048).

### Collinearity checks

Age and hospital were strongly collinear (point-biserial correlation = 0.76, p = 1.8×10□¹□), making it impossible to disentangle their respective contributions in subsequent models. Age was therefore retained as a continuous variable in preference to hospital.

Among DTI metrics, strong collinearity was observed between MD and AD (ρ = 0.90, p = 2.6×10□□□), MD and RD (ρ = 0.94, p = 4.0×10□□³), and AD and RD (ρ = 0.73, p = 8.5×10□²□). Since MD captures information from both AD and RD, only MD was retained in subsequent analyses.

### Model based on effective connectivity

Two equivalent models of D1 latency achieved significance (both p = 9.9×10□□, ΔBIC = 1.88) out of the 255 tested models: one including epilepsy type and stimulation site as predictors, and one including stimulation site alone. Post-hoc ANOVA identified robust effects of stimulation site in both models (p = 2.8×10□□ and p = 4.8×10□□ respectively) and of epilepsy type in the full model (p = 0.03). D1 latency was longer in TLE compared to ETLE (∼5.8 ms) and following hippocampal compared to amygdala stimulation (∼5 ms). Residual plots confirmed homoscedasticity. These results were confirmed in the cohort of 23 patients with favorable outcome (p = 2.1×10□^4^ and p = 8.9×10□^5^). Following hippocampal stimulation, D1 latency was significantly longer in TLE than in ETLE patients (∼5.6 ms, p = 0.03). No significant predictor was identified for D1 latency following amygdala stimulation.

None of the 255 candidate models tested reached significance for D1 amplitude, arguing against any robust predictor among those explored.

### Model based on mixed structural-effective connectivity

The BLMM incorporating both effective and structural connectivity was subsequently performed on D1 timing only (a post-hoc decision based on the lack of significant effective connectivity model for D1 amplitude), and converged satisfactorily across all four chains (RL < 1.01 for all parameters). The Bayesian R² was 0.47 (CI: 0.31 to 0.59).

Several predictors showed robust associations with D1 latency (Figure 3). Recordings within the EZ were associated with shorter D1 latency (β = -9.2 ms, CI: -16.4 to -2.2), with longer latency more associated with stimulation of the hippocampus compared to the amygdala (β = +7.2 ms, CI: 3.6 to 10.8), TLE compared to ETLE (β = +10.2 ms, CI: 3.6 to 16.8), and ipsilateral stimulation relative to the EZ compared to contralateral (β = +7.3 ms, CI: 1.6 to 13.0), indicating that D1 latency reflects both epileptogenic status and independent clinical/anatomical factors. Epilepsy duration, stimulation–recording Euclidean distance, amplitude, age, subtype, and tract presence showed no robust effect.

QA was positively associated with D1 latency (+0.50 ms per unit of QA, CI: 0.27 to 0.66), in contrast with FA which was negatively associated (−0.30 ms per unit of FA, CI: -0.49 to −0.06). Tract length and mean diffusivity showed no robust effect.

Significant DTI metrics (i.e., QA and FA) and CCEPs features (i.e., D1 timing) together demonstrated a significant added value to the null models including only significant DTI metrics (p=0.004) or CCEPs features (p<0.0002).

### Biomarker of EZ

The equation to obtain corrected latency and subsequent EZ score based on all significant variables extracted from the BLMM is

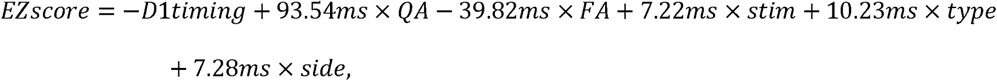

where D1timing (ms), QA and FA should be replaced by neurophysiology/neuroimaging values and stim by 1 for hippocampus stimulation and 0 for amygdala stimulation, type by 1 for TLE and 0 for ETLE, side by 1 for stimulation ipsilateral to the EZ and 0 for stimulation contralateral to the EZ (1 for both sides if bilateral EZ).

Using this equation in our cohort of patients who underwent both CCEPs and DTI, we obtained an AUC of 0.83 (p = 0.02, CI: 0.68 to 0.89) and the optimal threshold defined by Youden index was -13.55 ms (sensitivity 86%, specificity 71%). Coefficient estimates remained stable across folds after leave-one-out analyses (QA, 92.2±8.8ms; FA, -37.6±10.3ms; stim, 7.12±0.88ms; type, 10.12±1.27ms; side, 6.85±1.05ms), and pooled out-of-sample AUC was comparable to the in-sample estimate (0.82). Pooled sensitivity and specificity were less balanced (86% and 61%, respectively; overall accuracy 65%). A sensitivity analysis excluding the 3 folds with marginal Rhat deviations left the pooled AUC essentially unchanged (0.82), confirming that classification threshold instability across patients (rather than convergence issues) accounts for the imbalance between sensitivity and specificity (accuracy 75%, sensitivity 82%, specificity 73% in this subset, using a single pooled threshold rather than per-patient thresholds).

Nonetheless, the EZ score remains based on clinical data a priori not determined. Subsequently, the EZ probability was developed using an elastic net model (at λ_min = 0.0021) to free from any assumptions, which retained all three predictors, with QA and FA contributing the largest effects (QA_std = 2.760, FA_std = −2.782) and a smaller contribution from D1 timing (D1_timing_std = −0.052, intercept = −3.992), where QA_std, FA_std and D1_timing_std represent the standard deviation of each parameter.

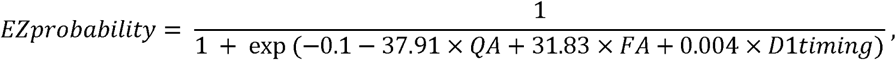

where D1timing (ms), QA and FA should be replaced by neurophysiology/neuroimaging values. In leave-one-patient-out cross-validation, pooled out-of-sample performance was: balanced accuracy 84% (CI: 63 to 96%), sensitivity 86%, specificity 82%, and overall accuracy 83% (CI: 60 to 98%), and final threshold was set at 0.28 ± 0.098, showing that neurophysiological and structural data alone can accurately identify epileptogenic contacts.

## Discussion

This study identified multiple determinants of D1 latency, spanning clinical characteristics, neurophysiological features, and white matter microstructure, with contributions from both physiological and pathological processes. The combination of these features supported the development of a multimodal biomarker of epileptogenicity in the amygdala-hippocampus complex.

### D1 timing as a marker of epileptogenicity

D1 timing - but not amplitude - was associated with epileptogenicity, even in a multivariate framework controlling for numerous covariables, consistent with previous works (7,9). However, the interpretation of shorter latency within the EZ warrants caution. In this cohort, epileptogenic contacts (−9.2 ms) necessarily co-occur with TLE (+10.2 ms) and ipsilateral stimulation (+7.3 ms), both of which independently increase latency. The net observed effect therefore reflects the balance between these opposing contributions rather than a straightforward facilitation effect, meaning that D1 latency would be:

- at its reference level (i.e., not delayed) in case of contralateral ETLE,
- delayed by 8.3 ms in case of ipsilateral mTLE (i.e., stimulation within the EZ; −9.2ms+10.2ms+7.3ms),
- delayed by 17.5 ms in case of ipsilateral TLE not involving the amygdala-hippocampus complex (+10.2ms+7.3ms),
- delayed by 10.2 ms in case of contralateral TLE regardless of amygdala-hippocampus involvement (+10.2ms), and
- delayed by 7.3 ms in case of ipsilateral ETLE (+7.3ms; Figure 4).

**Figure 4.**
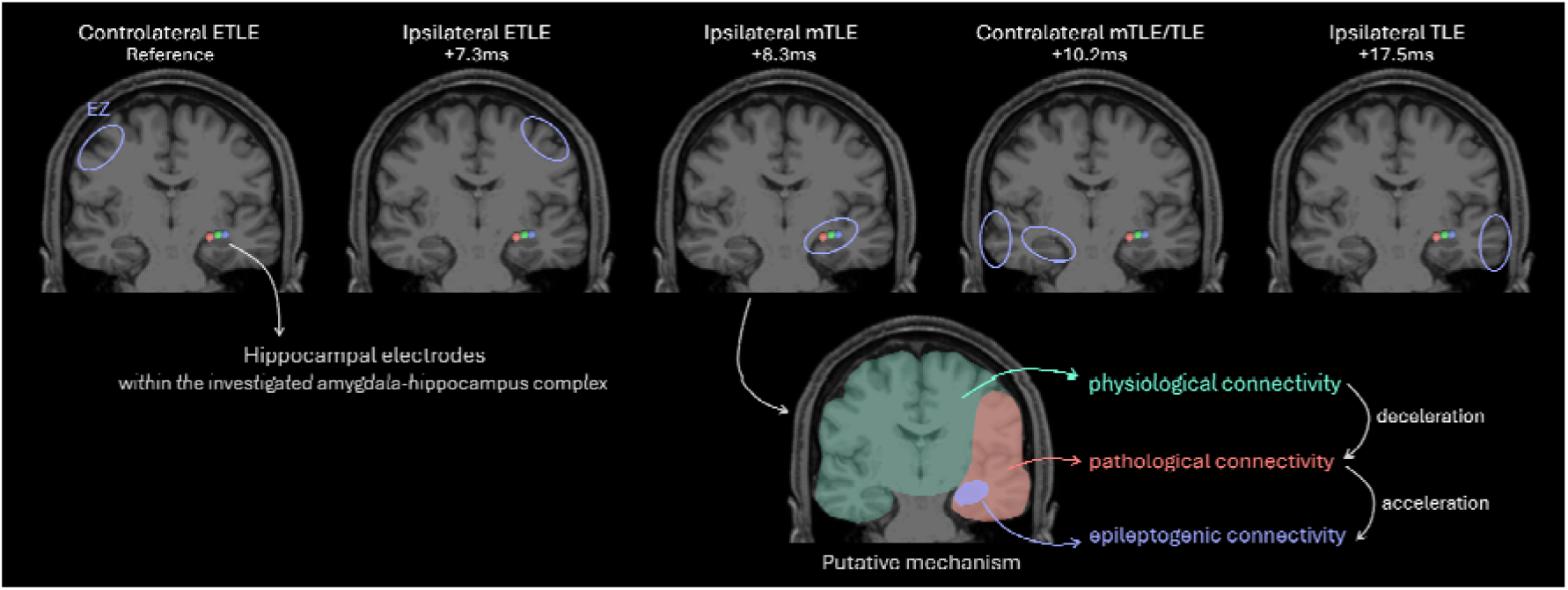
Latency of connectivity depending on the epileptogenic zone localization. Five examples of EZ localization (blue circle) are displayed on the top panel and average latencies of CCEPs in the amygdala-hippocampus complex (three hippocampal electrodes in red, green, blue) reported accordingly, with an increasing latency from left to right. Putative mechanism for the differential delay is schematically represented on the bottom panel in an example of medial temporal lobe epilepsy. For the sake of clarity, the area with pathological connectivity was represented in red. However, we hypothesized gradual pathological changes in the propagation speed, explaining lower deceleration in ipsilateral ETLE compared to contralateral mTLE/TLE and ipsilateral TLE. Brain MRI and SEEG implantation from a single patient were used for this figure, regardless of the actual EZ localization of this patient. The implantation involved other depth electrodes on the same coronal slice (radiological convention), removed for visualization purpose.

These findings do not support a pure increase in propagation velocity within the EZ. Furthermore, given the frequent recruitment of mesial temporal structures in non-mesial TLE (38) and the well-documented propagation to the contralateral hippocampus during temporal lobe seizures (39), the amygdala-hippocampus complex cannot be considered to underlie physiological connectivity in these cases, in contrast with contralateral ETLE, where it is not part of the seizure network. The TLE association with longer D1 latency, compared to ETLE, is consistent with the well-documented structural alterations of mesial temporal white matter in TLE (40).

Effective connectivity is therefore least delayed within the EZ itself, and progressively more delayed across the propagation network. Increased propagation speed within the epileptogenic network could reflect seizure-driven myelination changes proportional to recurrent electrical activity (41), consistent with the additional contribution of FA and QA to the model. Whether such microstructural alterations are a cause or a consequence of epilepsy, however, cannot be determined from cross-sectional data. The contribution of QA and FA is also supported by the known relationship of epilepsy with myelin content abnormalities (42) and congruent with the hypothesis of modified myelination of the epileptogenic network, leading to lower latency of CCEPs (9).

### Focus on D1 and its structural correlates

CCEP waveforms are complex and extend well beyond D1, reflecting polysynaptic and network-level dynamics. However, D1 is thought to represent the earliest, most direct cortico-cortical propagation (43), making it the component most likely to be captured by structural white matter metrics derived from DTI. This theoretical alignment justifies the focus on D1 in this study, even though a minority of electrode pairs showed CCEPs in the absence of detectable direct tracts, suggesting possible subcortical relay involvement in the early phase of the response (44). QA and FA both showed robust associations with D1 latency, though in opposite directions. While QA and FA reflect partially distinct microstructural properties of white matter, fiber packing density and intra-voxel directional reduction respectively (26), their opposing effects may reflect the heterogeneity of tracts within the amygdala-hippocampus complex (3). Importantly, the magnitude of these effects was small (less than 1ms for a full unit of FA/QA) relative to the typical range of white matter values. Surprisingly, tract length (and Euclidian distance) did not show a robust independent effect on D1 latency. This finding may appear counterintuitive, as conduction time is theoretically proportional to distance. However, tract length alone captures only one dimension of propagation: when combined with D1 latency, it allows estimation of propagation velocity along individual fascicles, which may prove more informative than either measure in isolation. More importantly, this effect might be hidden by stronger physiological (different brain areas) and pathological (connectivity changes in the EZ) effects. We tested this hypothesis in Supplementary Materials and confirmed by significant model with length explaining D1 timing after removal of stronger effects. This analysis also confirmed Euclidian distance as an imperfect proxy of tract length.

Human depth electrode studies have similarly demonstrated direct, asymmetrical connections between amygdala and hippocampus (45). This is supported by two results: (i) stimulation of the hippocampus yielded longer latencies than amygdala stimulation and (ii) fewer CCEPs were detected in the amygdala compared with those in the hippocampus.

i. This finding may initially appear counterintuitive, given that both structures projected onto the same recording site. This difference could partly reflect local circuit dynamics rather than differences in connection distance alone. Single-pulse intracranial stimulation is known to recruit not only excitatory principal cells but also fast and slow inhibitory populations, which shape the timing and waveform of the early evoked response (46) and such local inhibitory recruitment within the hippocampal circuit could plausibly delay the net excitatory response captured by D1.
ii. This may partly reflect the greater structural and functional heterogeneity of the amygdala, which comprises multiple nuclei with distinct connectivity profiles (3). Treating the amygdala as a single entity likely introduces unexplained variance, reducing the predictability of CCEPs features from this structure compared to the more functionally homogeneous hippocampus.

### Towards a multimodal biomarker of the epileptogenic zone

The EZ score derived from the combination of neurophysiological, clinical, and structural features yielded an AUC of 0.83, demonstrating a strong discriminative performance between epileptogenic and non-epileptogenic contacts within the amygdala-hippocampus complex. Its main strength lies in the principled integration of variables spanning distinct levels of analysis: two clinical features (epilepsy type and EZ side), a neurophysiological feature (D1 timing), an anatomical constraint (stimulation site), and microstructural white matter metrics (QA and FA). Unlike single-feature approaches, this multimodal integration captures complementary and largely non-redundant sources of information, as supported by the robust, independent effects identified in the BLMM. Critically, the score relies on variables that are either readily measurable during routine CCEP acquisition or already available from presurgical work-up, making it a practical, closed-form tool rather than a purely theoretical construct. The required clinical information may be deduced from invasive and non-invasive presurgical investigation: epilepsy type can be hypothesized from seizure semiology, scalp EEG, and neuroimaging, and lateralization can often be determined by interictal findings such as spikes or focal slow waves, without requiring seizure recording (47). This combination of strong discrimination and clinical feasibility positions the EZ score as a candidate tool to strengthen the localization of the EZ and improve markers of epileptogenicity. While this score might have clinical utility, e.g., to support the implantation of RNS leads within the hippocampus (48) or a selective amygdala-hippocampectomy (49), the added-value of its clinical use remains presumably low due to the need of a strong assumption regarding the type of epilepsy and the side of the EZ. For this reason, we developed the EZ probability relying on neurophysiological and neuroimaging data only (i.e., D1 latency, QA and FA). This new formula does not rely on any clinical assumption, subsequently increasing the added value to patients with multiple hypotheses regarding the localization of the EZ (e.g., with bilateral spikes or misleading semiology).

### Limitations

The main limitation of this study is the small number of patients with available diffusion MRI data (n=15), which constrains the statistical power and generalizability of the structural connectivity analyses and the biomarker evaluation. Additionally, the spatial sampling inherent to SEEG, although mitigated by the focus on a consistently implanted region, remains a source of bias. Also, visual detection of CCEPs, while more robust against false positives than automated approaches, introduces inter-rater variability that could affect reproducibility. This issue remains of low probability in the present study since the inter-observer agreement demonstrates no significant difference between two independent observers. On another note, the notion of physiological connectivity used throughout this discussion should be interpreted with caution, as all recordings were obtained from patients with epilepsy, and even contacts or regions outside the EZ may be affected by network-level or chronic effects of the disease. Moreover, we described the EZ as the area of primary organization of epileptic seizures, which might be considered as an approximation. Nonetheless, we confirmed these results in the cohort of patients with favorable postoperative outcome (Engel 1), which supports these results being representative of the epileptogenic zone and not only the seizure-onset zone. Finally, external validation of the EZ score in an independent cohort will be needed to confirm its generalizability and discriminative performance.

### Perspectives

Beyond the amygdala-hippocampus complex, extending this multimodal approach to other brain structures systematically explored during SEEG could progressively enable the construction of a normative and pathological atlas of CCEP latencies, integrating structural connectivity as a reference framework. Such an atlas would provide a principled basis for distinguishing physiological from pathological effective connectivity across brain regions and epilepsy types. Ultimately, this type of biomarker could reduce the length of iEEG recordings by alleviating the need for multiple seizure recordings.

## Conclusion

By combining effective and structural connectivity, this study identified key determinants of D1 latency and delivered a multimodal biomarker of the epileptogenic zone within the amygdala-hippocampus complex. These results argued for integrating multiple levels of analysis rather than relying on single connectivity features. Extending this framework to other brain regions could pave the way toward a structurally informed atlas of effective connectivity in epilepsy.

## Data Availability

All data produced in the present study are available upon reasonable request to the authors.

## Acknowledgements

O.F. received a research and training fellowship for clinicians from the American Epilepsy Society, a research fellowship from the International Federation of Clinical Neurophysiology and was supported by NIH grants R61-NS-125568 and R33-NS-125568.

M.Jo. was supported by NIH grants R61-NS-125568 and R33-NS-125568.

N.S. received NIH grant R00NS138680.

M.Ja. was supported by NIH grant F31NS14552901.

S.D. was supported by NIH grant R01-NS-116504.

J.M.S. was supported by NIH grant R01-NS-116504.

K.A.D. received support from NIH grants R01-NS-116504, R61-NS-125568 and R33-NS-125568.

E.C.C. received NIH grants K23 NS121401-01A1 and 1R01NS148413, and funding from the Burroughs Wellcome Fund.

## Conflicts of interest

Nothing to disclose.

## Supplementary Materials

### i. Linear mixed model

#### LMM based on neurophysiological data

Possible predictors included amplitude/timing of D1 (amplitude if outcome variable is timing and vice versa), epilepsy duration (i.e., difference between age at SEEG recording and age at epilepsy onset), Euclidean distance between stimulation and CCEPs, age at SEEG recording as continuous variables and type of epilepsy (TLE vs. ETLE), subtype of epilepsy (mTLE vs. other), epileptogenic recording area (Yes vs. No), stimulation site (hippocampus vs. amygdala), laterality of stimulation relative to the epileptogenic zone as categorical variables.

#### BLMM based on neurophysiological and neuroimaging data

The same predictors were used for the BLMM, and non-collinear neuroimaging data. If tract length was zero (indicating no detected tract), it was treated as a structural absence rather than a near-zero value: a binary variable was created to capture this information, and zero values of DTI metrics were replaced by missing values, preserving the continuous nature of these variables while distinguishing structural absence from true measurements.

Weakly informative priors were used for all parameters (Normal (0, 10) for fixed effects and intercept, Cauchy (0, 2) for the random effect standard deviation, and Normal (0, 1) for sub-model coefficients consistent with standardized predictors), following standard recommendations for Bayesian mixed models (36).

### ii. Complementary information between CCEPs and DTI

The significance of the added modality was assessed using a permutation-based likelihood-ratio test adapted from the Freedman-Lane approach for partial effects. Response residuals from the reduced model were permuted to generate null outcomes, and reduced and full models were refitted 5,000 times to obtain an empirical null distribution of the LRT statistic and the corresponding permutation p-value. Given the heuristic extension of this approach to binary mixed-effects models, the empirical null distribution was compared with the asymptotic chi-squared distribution for validation.

### iii. EZ biomarker

Model stability was further assessed using leave-one-patient-out cross-validation, restricted to patients with at least one DTI-available contact. At each iteration, the full BLMM was refit on all remaining patients, and the resulting coefficients and train-derived Youden threshold were applied to the held-out patient’s DTI-available contacts. Model convergence was checked for each fold (Rhat < 1.01); for folds showing only marginal deviations from this threshold, a sensitivity analysis was performed, excluding these folds to assess their impact on model performance. Classification metrics were pooled across all held-out patients.

### iv. Elastic net classifier

#### Results

Using the more conservative λ_1SE = 0.024, D1 timing was excluded entirely from the model (coefficient shrunk to 0), leaving QA and FA as the sole predictors (QA_std = 1.4, FA_std = −1.2; intercept = −2.3). The corresponding equation was

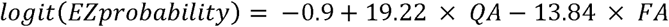

Pooled out-of-sample performance was balanced accuracy 0.78 (CI: 0.52 to 0.95), sensitivity 0.73, specificity 0.84, and overall accuracy 0.82 (CI: 0.61 to 0.97).

#### Discusssion

λ_1SE excluded D1 timing from the model, this is consistent with the elastic net’s conservative parsimony criterion rather than evidence against a CCEP contribution. The independent Freedman-Lane permutation test supports a significant partial effect of D1 timing when controlling for QA/FA, motivating our choice of λ_min as the primary reported model. The leave-one-out analysis excluded a potential overfitting.

### v. Null model using clinical information

A null model using only clinical variables available before CCEP/DTI acquisition was tested to estimate how much predictive value imaging data add. Because these are a few binary variables, only 7 possible combinations were observed in the data, and the resulting AUC would be based on too few discrete score values to be meaningfully interpreted as a curve. Among these clinical variables, only epilepsy type is independent of the EZ definition itself; at the patient level (Fisher’s exact test), it did not significantly discriminate EZ status (p = 0.106; 0/14 ETLE vs. 11/57 TLE patients had ≥1 EZ contact), supporting the added value of CCEP/DTI in the full model.

### vi. Distance/length effect

#### Methods

In a further secondary analysis restricted to hippocampus contacts (to remove effect from the brain area) contralateral to the EZ (to remove main pathological effect), we tested two separate linear mixed-effects models of D1 timing, with Euclidian distance between stimulating and recording electrodes (commonly used as a proxy of the length of pathways, not requiring DTI) or connection length (from DTI) as the fixed effect and a random intercept for subject (significance was assessed with a global F-test (p<0.05)).

#### Results

This subset included 81 observations from 11 patients. D1 timing was not significantly associated with distance (p = 0.283). In contrast, D1 timing was significantly associated with tracts length (p = 0.009), with longer connections associated with longer D1 latency. No significant heteroscedasticity was detected for either model.

## Notes

### Competing Interest Statement

The authors have declared no competing interest.

### Author Declarations

We retrospectively included all patients who underwent SPES protocols at Hospital of the University of Pennsylvania (HUP) and Children Hospital of Philadelphia (CHOP). At CHOP, SPES were performed under research protocol IRB15012226. At HUP, SPES were performed both as part of a research protocol under IRB821778 and as part of routine clinical care.

